# Development and Validation of a novel *in vitro* diagnostic test for endometriosis

**DOI:** 10.1101/2023.03.29.23287909

**Authors:** Bárbara Herranz-Blanco, Elza Daoud, Paola Viganò, Juan Antonio García-Velasco, Enrico Colli

## Abstract

**Objective:** To develop a non-invasive diagnostic test for endometriosis.

**Design:** We conducted two studies: the development study (study 1) aimed at (i) evaluating the ability of CA125, brain-derived neurotrophic factor (BDNF) and clinical variables in segregating between cases and controls and (ii) developing a diagnostic algorithm based on those results. In the validation study (study 2), the clinical performance of the developed *in vitro* diagnostic (IVD) test, in diagnosing endometriosis was validated.

**Interventions:** Serum samples and clinical variables extracted from psychometric questionnaires were collected from the Oxford Endometriosis CaRe Centre biobank (UK). Case/control classification was performed based on laparoscopy and histological verification of endometrial glands and/or stroma in the excised lesions.

**Main outcome measures:** Study 1 and 2 included *n*=204 and *n*=79 patients, respectively. CA125 and BDNF concentrations were determined using the endometriosis IVD ELISA kit. In study 2, serum CA125 and BDNF concentrations and clinical variables were introduced into the IVD test software hosting the data treatment algorithm, which generates the qualitative diagnostic result (“positive” or “negative”).

**Results:** Results from study 1 showed that, for both biomarkers levels, a statistically significant difference was found between cases and controls. Among all clinical variables related to patients’ medical history considered, six were significantly different between cases and controls: record of a previous surgery to investigate endometriosis presence, painful periods leading to referral for endometriosis presence, severity of menstrual pain during last cycle, age at first experience of intercourse pain, age at first regular use of painkillers and age at first diagnosis of ovarian cyst. In study 2, the novel endometriosis IVD test demonstrated sensitivity and specificity values of 46.2% (25.5-66.8%) and 100% (86.7-100%), respectively.

**Conclusion:** BDNF and CA125, together with patient’s clinical variables allowed efficient segregation between controls and endometriosis cases. Due to its high specificity, the novel endometriosis IVD test is an accurate rule-in non-invasive method, potentially contributing to diagnose endometriosis.

## Introduction

Endometriosis is an estrogen-dependent disease characterized by the growth of endometrial-like tissue outside the uterus (1,2). These lesions cause a chronic inflammatory reaction, which can lead to the generation of scar tissue and adhesions (3). Clinical symptoms include chronic pelvic pain, dysmenorrhea, and infertility (4). Endometriosis may increase a woman’s risk for chronic diseases such as cancer or autoimmune disorders and overall morbidity as well (5–7). Endometriotic lesions can occur at different locations, including the pelvic peritoneum and the ovary, or infiltrate pelvic structures below the peritoneal surface (deep endometriosis). According to these locations, three primary types of endometriosis have been defined: superficial peritoneal lesions (typically located on the pelvic organs or pelvic peritoneum), ovarian endometriomas and deep infiltrating endometriosis (DIE) (8).Endometriosis affects at least 10% of women of reproductive age and is associated with a high societal and economic burden: the average annual cost of healthcare and loss of productivity due to pain from endometriosis was $11,300 for affected women from the United States and nine European countries (9).

The clinical examination of symptomatic women does not reliably predict the presence of endometriosis. Imaging techniques often fail to diagnose the disease, mainly in early-stages or when only superficial peritoneal lesions are present. Imaging techniques are recognized to be useful in the identification of endometriomas, and in some cases of deep endometriosis.(1). Moreover, regardless of the lesion type and location, the interpretation of imaging findings is highly dependent of clinician’s experience and skills, which hinders diagnosis using those methods (10). Generally, women for whom there is high suspicion of endometriosis receive analgesics and hormonal medication without a prior definitive diagnosis (11). Diagnosing the disease in these cases becomes only possible via an invasive laparoscopy accompanied with histologic confirmation of lesions(10, 12). This may result in a delay in diagnosis ranging from 4 to 11 years on average between the first appearance of symptoms and the final confirmation of the diagnosis (13). In this context, the development of a non-invasive diagnostic tool is essential for faster diagnosis, appropriate treatment, and triaging potential surgical patients (14,15).

Multiple biomarkers have been studied as screening and triage tests for endometriosis (16,17). However, none of them has been implemented routinely in clinical practice (10). Out of all candidate biomarkers, Cancer Antigen 125 (CA125) has been extensively studied in endometriosis. CA125 is a high-molecular-weight glycoprotein expressed on the cell surface of some derivatives of embryonic coelomic epithelium, which are believed to be the precursors of endometriotic lesions (18). Studies found CA125 levels to be higher in patients with endometriosis, indicating that CA125 can be a useful marker for diagnosing endometriosis, distinguishing the severity of the disease, monitoring the effect of treatment and reflecting malignant transformation (19–21). A meta-analysis on the diagnostic accuracy of CA125 for endometriosis pooling 22 studies, including 3626 participants, showed that CA125 performed well as a rule-in test, but that a negative test result is unable to rule out endometriosis. In addition, the study showed that CA125 was significantly more sensitive for the diagnosis of moderate or severe endometriosis (stages III and IV) compared with minimal disease (22–24). Besides, CA 125 ≥ 30 U/m was highly predictive of endometriosis in women with symptoms of pain and/or subfertility, but CA 125□<30 U/ml was unable to rule out the disease (25).

Another interesting biomarker, brain-derived neurotrophic factor (BDNF), has been found to be linked with several pathways that are disturbed in women with endometriosis. BDNF is a neurotrophin (26) with a high-affinity to neurotrophic tyrosine receptor kinase 2 (NTRK2) also known as Tropomyosin receptor kinase B (TrKB), and this ligand-receptor pair participates in some aspects of uterine physiology (27). BDNF and NTRK2 were more significantly expressed in the uterus of women with endometriosis compared to disease-free controls (28); BDNF has been found to be dysregulated and overexpressed in ectopic but not in eutopic endometrial tissue. Interestingly, BDNF is a down-stream effector of estrogens mediating the pro-proliferative effects of estrogens promoting nociceptive pain (29,30). Estrogens strongly induce BDNF production by macrophages, and BDNF promotes neurogenesis through its binding to NTRK2 receptors on nerves. Release of pro-inflammatory mediators from mast cells, also triggered by estrogens, sensitizes peripheral nerve endings in endometriotic lesions, contributing to pain (31,32). Several research groups demonstrated that BDNF appears to be a good biomarker for early-stages (I-II) endometriosis (33–35).

With the aim of combining CA125, BDNF and clinical variables in order to develop a diagnostic tool that could identify all stages of endometriosis, two studies were conducted. The development study (study 1) aimed at (i) confirming the ability of BDNF, CA125 and patients’ clinical information to differentiate between cases and controls and (ii) developing a diagnostic algorithm based on the results. The validation study (study 2) aimed at establishing the clinical performance of the developed endometriosis IVD test, combining an ELISA kit to measure serum concentrations of BDNF and CA125 and a diagnostic software hosting the diagnostic algorithm.

## Methods

### Study populations

Studies were conducted on serum samples and data acquired from the Oxford Endometriosis CaRe Centre biobank (UK). This biobank emerged from the World Endometriosis Research Foundation (WERF) Endometriosis Phenome and Biobanking Harmonisation Project (EPHect) consensus on standardization and harmonization of phenotypic surgical/clinical data and biologic sample–collection methods in endometriosis research. Patients included in this biobank were of reproductive age (18 – 50 years old) and were undergoing a laparoscopy due to suspicion of endometriosis. This biobank comprised serum samples and patient’s clinical information (from questionnaires) collected before surgery along with surgical information collected during the procedure. Patients were classified as controls and cases and anonymized in the biobank. Patients were classified as cases if endometriosis was confirmed by laparoscopy and histological evaluation of excised lesions, and as controls if endometriosis lesions could not be visualized during the procedure or confirmed by laparoscopy and histology. The patients with endometriosis were classified in stages after laparoscopy according to the revised American Society of Reproductive Medicine (rASRM) classification. Also, endometriosis was classified per lesion location; superficial, endometrioma, and/or DIE depending on imaging and surgical findings. Endometriosis was classified as “superficial” if superficial endometriosis lesions only were found in the ovaries or in the peritoneal cavity. Endometriosis was classified as “endometrioma” if endometriomas were found in the ovaries with or without superficial endometriosis. Endometriosis was classified as “DIE” if infiltrative lesions were reported in the peritoneal cavity with or without the presence of superficial endometriosis. Endometriosis is classified as “endometrioma + DIE” if DIE was found in the peritoneal cavity along with endometriomas (with or without superficial endometriosis). For 5 patients only, this classification was not available.

The experimental protocols were approved by the Ethics committee of CEIm HM Hospitales. Two cohorts of patients were considered:

#### Development cohort

Serum samples from *n*=204 patients were included in the development study: n=136 patients with endometriosis and *n*=68 controls. Table 1 depicts the demographic characteristics of those patients. In this study, low- and high-stage endometriosis were equally represented in the cases group (stages I-II, 50% and stages III-IV, 50%).

**Table 1.** Demographic characteristics of the patients in the development cohort.

|  | **Controls**<br>**N=68** | **Cases**<br>**N=136** |
| --- | --- | --- |
| **Age years (mean ± SD)** | 33.5 (5.96) | 35.6 (6.42) |
| **BMI (mean ± SD)** | 25.38 (4.63) | 26.46 (5.32) |
| **rASRM classification** |  |  |
| **I–II** | - | 68 (50%) |
| **III–IV** | - | 68 (50%) |
| **Endometriosis Classification** |  |  |
| **Superficial** | - | 54 (39.7%) |
| **Endometrioma** | - | 26 (19.1%) |
| **DIE** | - | 29 (21.3%) |
| **DIE + endometrioma** | - | 25 (18.4%) |
| **Unclassified** | - | 2 (1.5%) |
| **Other gynaecological conditions** |  |  |
| Ovarian cysts | 28 | 66 |
| Ovarian cancer | 1 | 6 |
| Uterine fibroids | 7 | 25 |
| Adenomyosis | 0 | 7 |

Note. BMI= Body Mass Index; rASRM= revised American Society for Reproductive Medicine, DIE = Deep Infiltrative Endometriosis

#### External Validation cohort

Serum samples from *n*=79 patients were included in the validation study: n=52 patients with endometriosis and *n*=25 controls. Table 2 depicts the demographic characteristics of those patients. In this study, low stage (I-II) endometriosis patients represented 81% of the cases.

**Table 2.** Demographic characteristics of the patients in the external validation cohort.

|  | **Controls**<br>**N=25** | **Cases**<br>**N=52** |
| --- | --- | --- |
| **Age years (mean ± SD)** | 35 (6.44) | 35 (6.47) |
| **BMI (mean ± SD)** | 26 (5.23) | 26 (5.14) |
| **rASRM classification** |  |  |
| I–II | - | 42 (81%) |
| III–IV | - | 7 (13%) |
| Missing information |  | 3 (6%) |
| **Endometriosis Classification** |  |  |
| Superficial | - | 25 (48.1%) |
| Endometrioma | - | 3 (5.8%) |
| DIE | - | 14 (26.9%) |
| DIE + endometrioma | - | 8 (15.4%) |
| Unclassified | - | 3 (5.8%) |
| **Other conditions** |  |  |
| Ovarian cysts | 11 | 16 |
| Ovarian cancer | 0 | 4 |
| Uterine fibroids | 3 | 4 |
| Adenomyosis | 1 | 1 |

Note. BMI= Body Mass Index; rASRM= revised American Society for Reproductive Medicine, DIE = Deep Infiltrative Endometriosis.

**Table 3.** Performance characteristics of the IVD test in the development study.

| Model | Area Under Curve | Youden's index | Accuracy | Sensitivity | Specificity |
| --- | --- | --- | --- | --- | --- |
| At 95% specificity | 0.867<br>(0.819 – 0.915) | 47.1%<br>(37.3 - 56.8%) | 66.2%<br>(59.2 - 72.5%) | 51.5%<br>(42.8 - 60.1) | 95.65%<br>(86.8 - 98.9%) |
| At 95% sensitivity | 0.867<br>(0.819 – 0.915) | 44.1%<br>(31.7 - 56.5%) | 79.9%<br>(73.6 - 85%) | 95.6%<br>(90.2 - 98.2) | 48.5%<br>(36.4 - 60.9%) |
| At maximum Youden's index | 0.867<br>(0.819 – 0.915) | 58.8%<br>(46.7 - 70.9%) | 82.4%<br>(76.3 - 87.2%) | 88.2%<br>(81.3 - 92.9%) | 70.6%<br>(58.1 - 80.7%) |
| At maximum accuracy | 0.867<br>(0.819 – 0.915) | 58.1%<br>(45.9 - 70.3%) | 82.4%<br>(76.3 - 87.2%) | 89%<br>(82.2 - 93.5%) | 69.1%<br>(56.6 - 79.5%) |

### Sample collection

The specimens were collected and handled following the World Endometriosis Research Foundation Standard Operating procedures (Rahmioglu et al. 2014) after receiving patients’ consent. Patients were asked to fast for at least 10 hours prior to blood collection. Serum samples were stored in a biobank at -80 ºC for up to 5 years and were transferred to the laboratory analysis site.

### ELISA method: CA125 and BDNF concentrations

The IVD test ELISA (Enzyme-Linked Immunosorbent Assay) is a solid-phase sandwich enzyme-immunoassay for the quantitative determination of BDNF and CA125 in human serum. Each biomarker was determined in a different set of wells. The ELISA plate was coated with an antibody directed against either BDNF or CA125. BDNF or CA125 from samples and standards bind to the antibodies and were immobilized on the plate. Unbound biotin conjugate was washed off with washing solution. In a further step, streptavidin-HRP conjugate was added, and bound to the biotin. Unbound streptavidin-HRP was washed off with washing solution. Finally, a substrate solution was added, and the existing complex catalyzed the chemical reaction of the substrate into a colored chemical entity. The enzymatic color reaction was stopped after a defined period of time. The concentration of the colored chemical correlating proportionally to the concentration of the antibody was measured photometrically.

### Software input and score calculation

In the validation study only, upon collection of all the essential input parameters (serum CA125, serum BDNF and clinical variables), these data were introduced by the laboratory technicians into the IVD test diagnostic medical software’ hosting the data treatment algorithm. The algorithm outcomes were calculated and classified as positive or negative depending on whether the value was above or below the threshold value, respectively.

### Statistical analysis

Statistical analyses were performed using the software R, version 4.1.3 (R Foundation for Statistical Computing, Vienna, Austria), blinded to the surgical and imaging findings. Normal distribution was checked using the Shapiro-Wilk test. Because BDNF and CA125 levels did not follow a normal distribution, Mann-Whitney U analysis was used to compare BDNF and CA125 values between cases and controls. Sample sizes were chosen so that the 95% confidence interval does not exceed 0.3 for sensitivity and specificity outcomes around the expected value. To evaluate the importance of including both BDNF and CA125 in a diagnostic model, three logistic regression models with CA125 and BDNF as predictors were generated: one comparing the controls with all the cases, one comparing the controls with low-stage disease (S1-S2) and one comparing the controls with high-stage disease (S3-S4). Upon generation of these models, the Akaike information criterion (AIC) was applied during backward stepwise regression to identify whether or not BDNF and CA125 could identify endometriosis cases in the model.

Based on the results, CA125, BDNF and selected clinical variables were combined into a multivariable logistic regression model. Missing data were estimated by imputation: a threshold of 10% for each predictor was used as the maximum proportion of missing data for imputation. At each cut-off, sensitivity and specificity were computed together with the 95% confidence interval (CI). To compare the performance of the different regression models, we used ROC (Receiver Operating Characteristic) curves (Delacour et al., 2005). These allow comparison of specificity (proportion of negatives, i.e., controls, correctly identified as negatives) and sensitivity (proportion of positives, i.e. endometriosis cases, correctly identified as positives) of different models for different cut-off values. The higher the AUC (Area Under Curve) of these curves, the better the method. The maximum possible AUC is 1, which would indicate a perfect classifier. The Wilson score with continuity correction (Newcombe, 1998) was used to estimate 95% confidence intervals for accuracy, specificity, and sensitivity results. After selecting the most accurate model, the score was derived based on the final predictors and the corresponding regression coefficients. Rule-in cut-off and associated sensitivity were derived in the development cohort based on a specificity ≥90%.

In total, 122 clinical variables were considered for inclusion in the multivariable diagnostic algorithm. The predictors with a significant number of missing data points, with a significant correlation with age at time of surgery or a significant association with another candidate predictor (with a more significant association with endometriosis) were excluded for multivariable analysis. Chi-squared and Cochran-Armitage tests were used to determine which categorical variables were most strongly associated with endometriosis. Mann-Whitney U analysis was used for numerical variables.

In the validation study, algorithm scores and associated outcomes (positive diagnosis if the score was higher than the defined cut-off and negative diagnosis if the score was lower than the defined cut-off) were computed by the IVD test software. Based on these results, the primary (sensitivity and specificity) and secondary (accuracy, and AUC) performance parameters were calculated and reported, together with their 95% confidence intervals. The primary performance parameters results were compared with the values of the acceptance criteria established in the development study, to conclude whether the clinical performance of the device meets the criteria, i.e., whether the device can adequately classify the study subjects as positive or negative for endometriosis. In concrete, the sensitivity and specificity in the validation study should not be lower than the lower limits of the sensitivity and specificity 95% confidence intervals in the algorithm development study. Because the prevalence of stage I-II in the validation study was significantly higher than in the development study (Chi-square= 18.06, p<0.001)., the outcomes in the validation were weighted to give equal representation to the low-stage and high-stage groups.

## Results

### Diagnostic performance of CA125 and BDNF in endometriosis

Figure 1 displays BDNF and CA125 values in cases and controls. The Mann-Whitney U analysis showed that both BDNF and CA125 were significantly higher in cases than in controls (*p* < 0.01 and p<0.001, respectively).

**Figure 1.**
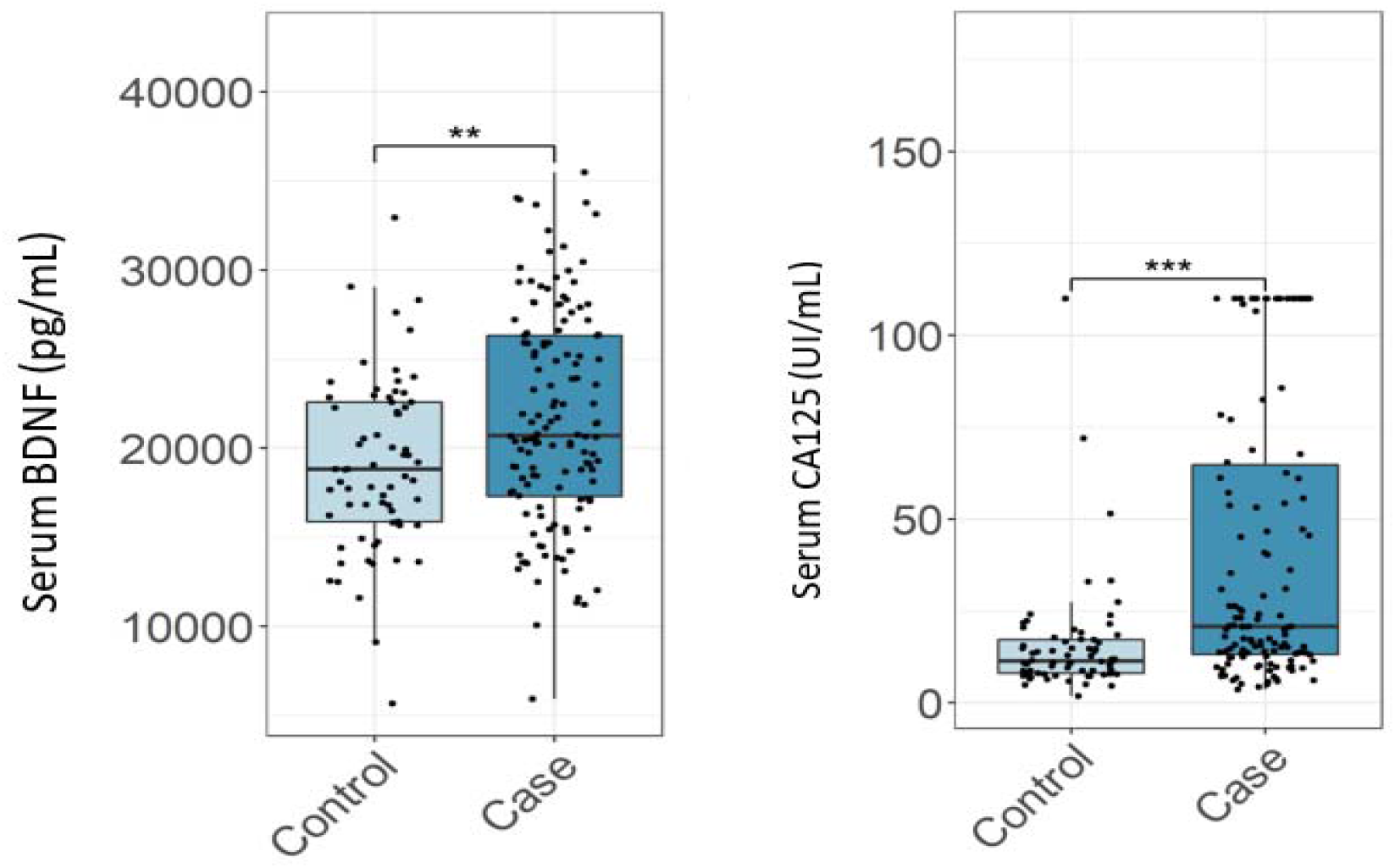
Serum concentration of CA125 and BDNF in endometriosis patients and controls. Asterisk signs above boxplots indicate a statistically significant difference in median value between the indicated population (** : p < 0.01; *** : p < 0.001) as established by a Mann-Whitney U test.

In backwards stepwise regression analysis based on AIC, for the comparisons of the control group with all cases and with the high-stage disease cases, both CA125 and BDNF were retained, meaning they were both independently informative as predictors of endometriosis. For the comparison of the control group with low-stage disease, only BDNF was retained, meaning that only BDNF was independently informative as predictor of low-stage endometriosis.

Taken together, both CA125 and BDNF are able to distinguish controls from endometriosis cases, with the former performing very well in the high-stage group and the latter the better contributor for the low-stage group. Therefore, both parameters were justified for inclusion in a multivariable model for endometriosis diagnosis.

### Development of prediction model for endometriosis

Among all clinical variables related to patients’ medical history that were considered, chi-square analysis showed that only three qualitative variables were significantly different between cases and controls. Most significantly, a previous history of surgery to examine endometriosis (even if the disease was not diagnosed) was more common among patients tested positive for current endometriosis (54.6% in cases, 10.6% in controls, *p* < 0.001). Painful periods as a symptom leading to a referral for endometriosis was also associated with a positive diagnosis with strong statistical significance (76.4% in cases, 36.8% in controls, *p* < 0.001). Other significant variables was the severity of the last menstrual cycle pain, with moderate/severe pain being more frequent in cases than in controls (78.0% in cases, 47.1% in controls, *p* < 0.01). For numerical (quantitative) variables, the median of three of them were significantly different between cases and controls in the Mann-Whitney U test: age at first regular use of painkiller (*U* = 343.5, *p* = 0.038), age at first diagnosis of ovarian cyst (*U* = 334.5, *p* = 0.023), and age at first experience of intercourse pain (*U* = 1201, *p* = 0.009).

In the final revised model, the eight above discussed variables were considered:CA125, BDNF and the six clinical variables, i.e. record of a previous surgery to examine for endometriosis, painful periods as a symptom leading to referral for endometriosis, severity of menstrual pain during last cycle, age at first experience of intercourse pain, age at first regular use of painkillers and age at first diagnosis of ovarian cyst.

To estimate its performance on independent data, a logistic regression model was repeatedly generated on 80% of the data and evaluated on the remaining 20%. The final model, generated from all algorithm development data, was optimized for high specificity to render a rule-in test with a low rate of false positives (36). This model has an AUC of 0.867 with a sensitivity of 51.5% at a specificity of 95.6%.

### Clinical performance evaluation (validation of the IVD test)

The diagnostic performance of the IVD test, comprising the ELISA kit method for the determination of BDNF and CA125 together with the diagnostic algorithm established in the development study was evaluated in an independent sample cohort. The endometriosis IVD test had a sensitivity (after weighing for disease stages) of 46.2% (95% CI: 25.5-66.8%) and a specificity of 100% (95% CI: 86.7-100%). The accuracy was 64.1% (95% CI: 50.4-77.8%) and the AUC was 0.758 (95% CI: 0.650-0.867). With an observed diagnostic specificity in this clinical performance study of 100%, the target specificity of 86.8% (or higher) is met. A good specificity was the primary objective because this assay is primarily intended to aid in identifying individuals with endometriosis. For the sensitivity, a mid-range sensitivity rather than a low sensitivity was desired to ensure that a significant proportion of the test population will test positive.

## Differential diagnosis

### Confounding conditions

Further, we investigated whether other gynaecological conditions could interfere with the performance of the IVD test, rendering a positive test result when endometriosis is not present (false positive). The conditions considered as potentially confounding were non endometriosis benign ovarian cysts, ovarian cancer, uterine fibroids and adenomyosis. In this respect, out of 93 controls included in the development (*n*=68) and validation (*n*=25) studies, 42% (*n*=39) had other ovarian cysts and 11% (*n*= 10) had uterine fibroids. Only 2 patients (2%) had a positive test result (false positive), and thus the effect of these potentially confounding conditions on the test is considered very limited.

### Detection of superficial endometriosis

We have analysed the capacity of the endometriosis IVD test to identify the cases presenting just with superficial endometriosis. In the development cohort, endometriosis could be classified into different groups (into superficial endometriosis, endometrioma, endometrioma + DIE and DIE) for 134 out of 136 cases. In the validation cohort, the classification could be done for 50 out of 53 cases. Out of those *n*=184 patients in total (both cohorts), *n*=79 patients had superficial endometriosis (43%). With the endometriosis IVD Test, *n*=25 of the *n*=79 (32%) cases with superficial endometriosis were detected.

## 4. Discussion

We developed a non-invasive in vitro diagnostic (IVD) test for endometriosis using a step-by-step approach. In the development study, the ability of BDNF and CA125 to differentiate between cases and controls was confirmed. Based on those results, the IVD test, consisting of an ELISA kit for the determination of serum concentrations of BDNF and CA125 and a data treatment algorithm hosted in a diagnostic medical software was developed. In the validation study, the clinical performance of The IVD test in diagnosing endometriosis was established. Main results are discussed below.

First, although no cut-off values were found, CA125 and BDNF levels were demonstrated to be elevated in patients with endometriosis; with CA125 mostly able to identify high-stage endometriosis and BDNF performing well for both low- and high-stage disease. This confirms what was previously found by other research groups: BDNF concentrations are higher in endometriosis patients than in controls in plasma (34,37,38) and serum (33,35,39). We chose to measure BDNF concentration in serum because, as previously shown, during centrifugation, all of the BDNF content is released from platelets, reducing measurement errors related to blood handling, storage, and analysis encountered with plasma samples. (40,41). Although there is much confounding evidence on the validity of CA125 as a biomarker for endometriosis, two meta-analyses showed that it could be used in conjunction with clinical information (21,24).

A number of controls in both development and validation studies had other gynecological conditions that could elevate the CA125 concentration in serum (e.g., benign ovarian cysts, uterine fibroids, ovarian cancer and adenomyosis) (23) and had a negative diagnosis (classified as true negative) using the IVD test. The presence of such confounding factors did not lead to any false positive result in the validation study. This is likely because the IVD test does not rely solely on CA125 but also on BDNF and the patient’s clinical information.

In the validation study, as the algorithm was optimized for specificity during development, the novel endometriosis IVD test showed a limited sensitivity (46.2%) but a very high specificity of 100%, making it an excellent rule-in test able to minimize the risk of false positives. A rule-in test is considered the most appropriate approach given the chronic and non-life-threatening nature of the disease. A positive test result would aid the clinician in the diagnosis, when considered together with other clinical information. The diagnosis of women presenting only with superficial lesions by a non-invasive test is of special interest due to the limited value of existing imaging techniques for their identification (1,10), possibly leading to numerous misdiagnoses. Considering that the endometriosis IVD test was able to detect 32% of cases presenting with superficial lesions recruited in the studies, this diagnostic tool can provide an added value for the diagnosis of this type of disease. When the test is negative, the clinician may consider other causes for the symptoms or symptomatic treatment for pain, according to their usual practice. If the suspicion of endometriosis persists after a follow up consultation, the women can be re-tested at the discretion of the clinician. Our diagnostic test compares well with other benchmark diagnostic tests, such as prostate-specific antigen (PSA) to detect prostate cancer which has a sensitivity of 93% (95% CI 88%, 96%) and a specificity of 20% (95% CI 12%, 33%) (42).

An essential strength of this study is that all the participants underwent laparoscopy (gold standard diagnosis), a necessary component of algorithm development to provide the true clinical state of each participant. The diagnostic algorithm was developed based on *n*=204 patients in the development cohort. A total of 8 predictors were included in the multivariate logistic regression model: CA125, BDNF, record of previous surgery for endometriosis, painful periods leading to referral for endometriosis, age at first intercourse pain, age at first painkillers use, age at first ovarian cyst symptom and severity of menstrual pain during last cycle. After performing the IVD ELISA test, laboratory technicians can introduce CA125 and BDNF results in a diagnostic medical software along with patients’ medical information. The software hosting the algorithm calculates a score, which according to a certain cutoff value, provides a diagnosis.

This novel endometriosis IVD test is a medical device that has been CE marked under the IVD Directive 98/79/EC. The test could be included in early workup to aid clinicians in the diagnosis of endometriosis when the disease is suspected, in conjunction with other clinical information, in order to facilitate timely access to a correct disease management.

## 5. Conclusion

We have developed and validated a non-invasive *in vitro* diagnostic test for endometriosis. The excellent rule-in performance of this test could provide a significant value in the clinical management of this disease.

## Data Availability

The subjects in this trial have not concomitantly been involved in other randomized trials.
Data regarding any of the subjects in the study has not been previously published unless specified.
Data will be made available to the editors of the journal for review or query upon request.

## Bibliography

1. Zondervan KT, Becker CM, Missmer SA. Endometriosis. New England Journal of Medicine 2020;382(13):1244–56.

2. Zondervan KT, Becker CM, Koga K, Missmer SA, Taylor RN, Viganò P. Endometriosis. Nat Rev Dis Primers 2018;4(1):1–25.

3. Symons LK, Miller JE, Kay VR, Marks RM, Liblik K, Koti M, et al. The Immunopathophysiology of Endometriosis. Trends in Molecular Medicine 2018;24(9):748–62.

4. Taylor HS, Kotlyar AM, Flores VA. Endometriosis is a chronic systemic disease: clinical challenges and novel innovations. The Lancet 2021;397(10276):839–52.

5. Rossi H-R, Uimari O, Terho A, Pesonen P, Koivurova S, Piltonen T. Increased overall morbidity in women with endometriosis: a population-based follow-up study until age 50. Fertility and Sterility 2023;119(1):89–98.

6. Sinaii N, Cleary SD, Ballweg ML, Nieman LK, Stratton P. High rates of autoimmune and endocrine disorders, fibromyalgia, chronic fatigue syndrome and atopic diseases among women with endometriosis: a survey analysis. Hum Reprod 2002;17(10):2715–24.

7. Somigliana E, Vigano’ P, Parazzini F, Stoppelli S, Giambattista E, Vercellini P. Association between endometriosis and cancer: a comprehensive review and a critical analysis of clinical and epidemiological evidence. Gynecol Oncol 2006;101(2):331–41.

8. Simoens S, Dunselman G, Dirksen C, Hummelshoj L, Bokor A, Brandes I, et al. The burden of endometriosis: costs and quality of life of women with endometriosis and treated in referral centres. Human Reproduction 2012;27(5):1292–9.

9. Agarwal SK, Chapron C, Giudice LC, Laufer MR, Leyland N, Missmer SA, et al. Clinical diagnosis of endometriosis: a call to action. Am J Obstet Gynecol 2019;220(4):354.e1-354.e12.

10. Becker CM, Bokor A, Heikinheimo O, Horne A, Jansen F, Kiesel L, et al. ESHRE guideline: endometriosis. Hum Reprod Open 2022;2022(2):hoac009.

11. Dunselman G a. J, Vermeulen N, Becker C, Calhaz-Jorge C, D’Hooghe T, De Bie B, et al. ESHRE guideline: management of women with endometriosis. Hum Reprod 2014;29(3):400–12.

12. Jenkins TR, Liu CY, White J. Does response to hormonal therapy predict presence or absence of endometriosis? J Minim Invasive Gynecol 2008;15(1):82–6.

13. Ahn SH, Singh V, Tayade C. Biomarkers in endometriosis: challenges and opportunities. Fertility and Sterility 2017;107(3):523–32.

14. Irungu S, Mavrelos D, Worthington J, Blyuss O, Saridogan E, Timms JF. Discovery of non-invasive biomarkers for the diagnosis of endometriosis. Clin Proteomics 2019;16:14.

15. Nisenblat V, Bossuyt PMM, Farquhar C, Johnson N, Hull ML. Imaging modalities for the non-invasive diagnosis of endometriosis. Cochrane Database Syst Rev 2016;2(2):CD009591.

16. Nisenblat V, Bossuyt PM, Shaikh R, Farquhar C, Jordan V, Scheffers CS, et al. Blood biomarkers for the non-invasive diagnosis of endometriosis. Cochrane Database of Systematic Reviews [Internet] 2016 [cited 2023 Jan 26];(5). Available from: https://www.cochranelibrary.com/cdsr/doi/10.1002/14651858.CD012179/full

17. Barbieri RL, Niloff JM, Bast RC, Schaetzl E, Kistner RW, Knapp RC. Elevated serum concentrations of CA-125 in patients with advanced endometriosis. Fertility and Sterility 1986;45(5):630–4.

18. Chen Y, Pan M, Zuo Y, Yang B, Wang S. Research progress of CA125 in endometriosis: Teaching an old dog new tricks. Gynecology and Obstetrics Clinical Medicine 2022;2(4):191–8.

19. Nagamani M, Kelver ME, Smith ER. CA 125 levels in monitoring therapy for endometriosis and in prediction of recurrence. Int J Fertil 1992;37(4):227–31.

20. Shen A, Xu S, Ma Y, Guo H, Li C, Yang C, et al. Diagnostic value of serum CA125, CA19-9 and CA15-3 in endometriosis: A meta-analysis. J Int Med Res 2015;43(5):599–609.

21. Kafali H, Artuc H, Demir N. Use of CA125 fluctuation during the menstrual cycle as a tool in the clinical diagnosis of endometriosis; a preliminary report. European Journal of Obstetrics & Gynecology and Reproductive Biology 2004;116(1):85–8.

22. Karimi-Zarchi M, Dehshiri-Zadeh N, Sekhavat L, Nosouhi F. Correlation of CA-125 serum level and clinico-pathological characteristic of patients with endometriosis. Int J Reprod Biomed 2016;14(11):713–8.

23. Hirsch M, Duffy J, Davis C, Nieves Plana M, Khan K, on behalf of the International Collaboration to Harmonise Outcomes and Measures for Endometriosis. Diagnostic accuracy of cancer antigen 125 for endometriosis: a systematic review and meta-analysis. BJOG: An International Journal of Obstetrics & Gynaecology 2016;123(11):1761–8.

24. Hirsch M, Duffy JMN, Deguara CS, Davis CJ, Khan KS. Diagnostic accuracy of Cancer Antigen 125 (CA125) for endometriosis in symptomatic women: A multi-center study. European Journal of Obstetrics & Gynecology and Reproductive Biology 2017;210:102–7.

25. Chao MV. Neurotrophins and their receptors: A convergence point for many signalling pathways. Nat Rev Neurosci 2003;4(4):299–309.

26. Wessels JM, Wu L, Leyland NA, Wang H, Foster WG. The brain-uterus connection: brain derived neurotrophic factor (BDNF) and its receptor (Ntrk2) are conserved in the mammalian uterus. PLoS One 2014;9(4):e94036.

27. Wang S, Duan H, Li B, Hong W, Li X, Wang Y, et al. BDNF and TrKB expression levels in patients with endometriosis and their associations with dysmenorrhoea. Journal of Ovarian Research 2022;15(1):35.

28. Lim W, Bae H, Bazer FW, Song G. Brain-derived neurotrophic factor improves proliferation of endometrial epithelial cells by inhibition of endoplasmic reticulum stress during early pregnancy. Journal of Cellular Physiology 2017;232(12):3641–51.

29. Dong F, Zhang Q, Kong W, Chen J, Ma J, Wang L, et al. Regulation of endometrial cell proliferation by estrogen-induced BDNF signaling pathway. Gynecological Endocrinology 2017;33(6):485–9.

30. Godin SK, Wagner J, Huang P, Bree D. The role of peripheral nerve signaling in endometriosis. FASEB BioAdvances 2021;3(10):802–13.

31. Greaves E, Temp J, Esnal-Zufiurre A, Mechsner S, Horne AW, Saunders PTK. Estradiol Is a Critical Mediator of Macrophage-Nerve Cross Talk in Peritoneal Endometriosis. The American Journal of Pathology 2015;185(8):2286–97.

32. Perricos A, Ashjaei K, Husslein H, Proestling K, Kuessel L, Obwegeser R, et al. Increased serum levels of mBDNF in women with minimal and mild endometriosis have no predictive power for the disease. Exp Biol Med (Maywood) 2018;243(1):50–6.

33. Wessels JM, Kay VR, Leyland NA, Agarwal SK, Foster WG. Assessing brain-derived neurotrophic factor as a novel clinical marker of endometriosis. Fertil Steril 2016;105(1):119-128.e1-5.

34. Ding S, Zhu T, Tian Y, Xu P, Chen Z, Huang X, et al. Role of Brain-Derived Neurotrophic Factor in Endometriosis Pain. Reprod Sci 2018;25(7):1045–57.

35. Trevethan R. Sensitivity, Specificity, and Predictive Values: Foundations, Pliabilities, and Pitfalls in Research and Practice. Frontiers in Public Health [Internet] 2017 [cited 2023 Mar 6];5. Available from: https://www.frontiersin.org/articles/10.3389/fpubh.2017.00307

36. Giannini A, Bucci F, Luisi S, Cela V, Pluchino N, Merlini S, et al. Brain-Derived Neurotrophic Factor in Plasma of Women with Endometriosis. Journal of Endometriosis 2010;2(3):144–50.

37. Rocha AL, Vieira EL, Ferreira MC, Maia LM, Teixeira AL, Reis FM. Plasma brain-derived neurotrophic factor in women with pelvic pain: a potential biomarker for endometriosis? Biomark Med 2017;11(4):313–7.

38. Liang YF, Huang XM, Wen LL, Kang H, Tao MH, Ye MZ. [Relationship between serum brain-derived neurotrophic factor and clinical stage and dysmenorrhoea of enodmetriosis]. Zhonghua Yi Xue Za Zhi 2020;100(10):771–4.

39. Polacchini A, Metelli G, Francavilla R, Baj G, Florean M, Mascaretti LG, et al. A method for reproducible measurements of serum BDNF: comparison of the performance of six commercial assays. Sci Rep 2015;5:17989.

40. Polyakova M, Schlögl H, Sacher J, Schmidt-Kassow M, Kaiser J, Stumvoll M, et al. Stability of BDNF in Human Samples Stored Up to 6 Months and Correlations of Serum and EDTA-Plasma Concentrations. Int J Mol Sci 2017;18(6):1189.

41. Merriel SWD, Pocock L, Gilbert E, Creavin S, Walter FM, Spencer A, et al. Systematic review and meta-analysis of the diagnostic accuracy of prostate-specific antigen (PSA) for the detection of prostate cancer in symptomatic patients. BMC Medicine 2022;20(1):54.

